# Digital Health-Enabled Community-Centered Care (D-CCC): A Scalable Model to Empower Future Community Health Workers utilizing Human-in-the-Loop AI

**DOI:** 10.1101/2021.03.03.21252873

**Authors:** Sarah M. Rodrigues, Anil Kanduri, Adeline M. Nyamathi, Nikil Dutt, Pramod P. Khargonekar, Amir M. Rahmani

## Abstract

Digital Health-Enabled Community-Centered Care (D-CCC) represents a pioneering vision for the future of community-centered care. Utilizing an artificial intelligence-enabled closed-loop digital health platform designed for, and with, community health workers, D-CCC enables timely and individualized delivery of interventions by community health workers to the communities they serve. D-CCC has the potential to transform the current landscape of manual, episodic and restricted community health worker-delivered care and services into an expanded, digitally interconnected and collaborative community-centered health and social care ecosystem which centers around a digitally empowered community health workforce of the future.

## 1. Introduction

Recent global health trends necessitate a shift away from a patient-centered medical care system to an ‘upstream’ health promotion approach which meets the health and social needs of individuals in the communities where they live ^1^. A community-centered health and social care ecosystem, supported by a robust community health worker (CHW) workforce, aligns with this paradigm shift ^1^. Several states in the United States (US) have launched CHW initiatives as an alternative assistance to registered health care professionals to extend the reach of home delivered services ^2^. Further, recent data from the California Employment Development Department projects that an additional 200,000 CHWs will be needed by 2024 ^3^. A *rapid expansion of the CHW footprint is needed* to prevent unmet needs from escalating into expensive medical crises, particularly in vulnerable communities ^4^.

However, the current community-centered care (CCC) model suffers from an inability to scale in a cost-effective manner while critically preserving and/or expanding contextual and individually tailored approaches which are key to CHW-delivered care. The increasing use of digital technologies by CHWs within the current CCC model has the potential to expand the CHW footprint; however, existing digital technologies remain ad hoc and lack integration. We believe that an integrated, intelligent and automated closed-loop platform designed for, and with, CHWs has tremendous potential for improving upon the current CCC model by maximally amplifying and expanding the CHW footprint. This novel artificial intelligence (AI)-enabled closed-loop digital health platform has the potential to transform the current state of manual, episodic and restricted work to an expanded, digitally interconnected and empowered CHW workforce of the future. Our vision is a new model for CCC: Digital Health-Enabled Community-Centered Care (D-CCC). D-CCC integrates future digital health technologies, the future CHW workforce and the future health and social care needs of communities to connect the most vulnerable individuals within communities to critical health-related information, resources and services.

## 2. Community Health Workers

Community health workers (CHWs) are lay members of the community whose in-depth understanding of community culture and language uniquely positions them to provide culturally appropriate health-related services and ongoing behavioral support to the community ^5^. Through their shared lived experience and deep familiarity with social networks and community resources, CHWs serve as vital ‘bridges’ between vulnerable communities and health systems, critically connecting vulnerable individuals to information, resources and services ^5,6^

CHWs work in a variety of settings, from non-profit community-based organizations to government agencies and health care systems ^6,7^, and may range from formal, salaried employees to informal, volunteer-based community educators ^8,9^. As the activities and roles of CHWs are tailored to the unique health needs of the communities they serve ^10^, CHWs may operate under a diverse set of titles, including *promotor(a)s*, community lay workers, health navigators, outreach educators, peer support workers, and home visitors, among others. In this paper we use ‘CHW’ as an umbrella term intended to broadly capture the rich diversity of roles and titles held by this vital frontline lay health workforce ^5^.

As trusted members of the community, CHWs are critically positioned to support vulnerable patient populations ^11^ and serve as frontline agents of change by helping to reduce health disparities in underserved communities ^12^. Documented positive impacts of CHWs include health promotion, improved patient engagement, support with adherence to treatment, improved referrals and access to care, financial return on investment, and improved quality of care and health outcomes ^6^. CHW-delivered socio-behavioral interventions have demonstrated efficacy in improving health outcomes in chronic and non-communicable disease (NCD) care and management ^13^, including cancer ^14^, diabetes ^15–21^, asthma ^22,23^, cardiovascular disease ^24^, multiple medical comorbidities ^25^ and mental health ^26,27^. CHW-delivered interventions have also demonstrated efficacy in reducing (re-)hospitalization rates ^25,28–30^. Finally, CHWs are powerful drivers of decreased health care costs, particularly among patients with high starting health care costs, as well as underserved and minority populations ^13,15,31,32^. The efficacy of CHWs is grounded in their close connection to the community, ability to influence patient behaviors, and effective interaction with the larger health care team ^32–34^.

Recent global health trends, including the growing burden of chronic and NCDs, a focus on addressing the social determinants of health and reducing health disparities, and lessons learned from the COVID-19 pandemic, are necessitating a shift away from a patient-centered medical care system to a focus on ‘upstream’ health promotion through addressing the health and social needs of individuals in the communities where they live ^1^. In place of the traditionally siloed existence of medicine, public health, and mental health ^35^, a community-centered health and social care ecosystem, *supported by a robust CHW workforce*, will be critical to realizing this needed paradigm shift ^1^. A *rapid expansion of the CHW footprint is needed* to prevent unmet needs from escalating into expensive medical crises, particularly in vulnerable communities ^4^.

## 3. The Current Use of Digital Technologies in Community-Centered Care (CCC)

CHWs typically operate within a community-centered care (CCC) model, as shown in Figure 1. Under this model, a smaller number of registered health care professionals (e.g., registered nurses, social workers) supervise a larger number of CHWs who in turn provide culturally tailored, language-appropriate, and individualized care for a yet larger number of clients. While this pyramidal care model has demonstrated improved health outcomes and cost savings ^25,30^, the current CCC model suffers from an inability to scale in a cost-effective manner while preserving (or expanding) the individually-tailored quality care provided by CHWs. However, incorporation of digital health technologies into the CCC model is demonstrating potential for amplification and expansion of the CHW footprint while critically preserving the community-specific contextual expertise and individualized care delivered by CHWs.

**Figure 1:**
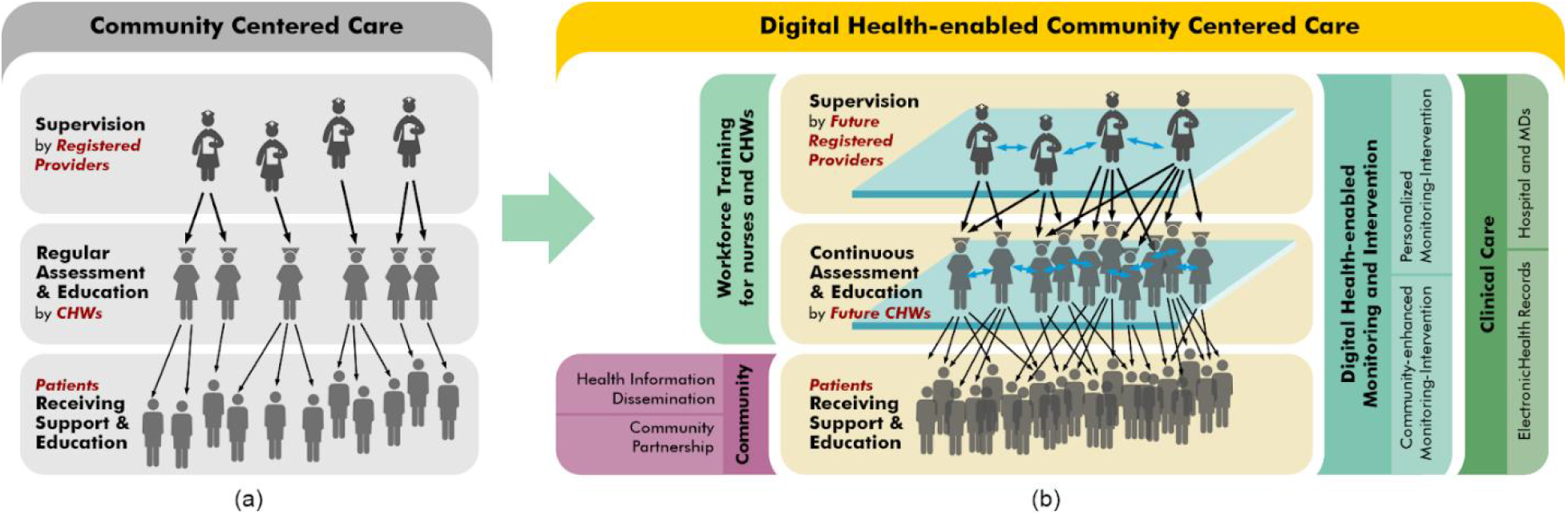
(a) The current CCC model vs. (b) our proposed D-CCC model.

### 3.1 Current Digital Technologies Utilized in CCC

Use of digital technologies has recently been increasing among CCC organizations and is demonstrating potential to enhance CHWs’ reach and diffusion of health information within communities ^5,36,37^. A recent scoping review ^5^ of the use of mHealth technologies and interventions among CHWs globally highlighted key benefits and challenges identified in the literature. Key benefits included promotion of health equity, reduced time to diagnosis, extension of health information and services to diverse areas, improved adherence to treatment plans, increased self-efficacy of patients and CHWs, and improved attitudes of, and toward, CHWs and their role ^5^. Key challenges included a lack of evaluation of mHealth outcomes, development of mHealth tools and apps without cultural relevance, lack of access to, and knowledge of, mobile technologies within communities, need for effective training for CHWs to adopt mHealth tools and the need for improved communication between and among health care teams working with CHWs ^5^. Similarly, in a recent narrative review of the literature, Mishra et al. (2019) ^10^ identified three benefits and three challenges to incorporating digitalization into CHW practice. Benefits included improved access and quality of services, increased efficiency in training and personnel management, and leveraging of data generated across grass-roots platforms to further research and evaluation ^10^. Challenges included funding for CHW programs, digital health literacy of CHWs, and systemic challenges related to motivating CHWs, including adequate CHW supervision ^10^.

Digital platforms currently used by CHWs and CCC organizations include mobile-based networking devices, web-applications, videoconference and mobile applications ^10^. CHWs currently use mobile phones, tablets and other digital devices ^38^ in a task-specific manner (e.g., digital blood pressure monitoring devices, glucometers and spirometers) ^10,39^. Digital alerts, reminders, notifications, checklists and decision-support tools facilitate compliance to protocols and improve the quality of care provided by CHWs ^10,40^. Whereas the continuous monitoring of physiological parameters (e.g., heart rate, blood pressure) has typically been available only in hospital settings due to the need for special equipment and medical expertise, the development of relatively low-cost, non-intrusive sensors are presenting opportunities to enable ubiquitous monitoring of clients’ health and wellbeing. Current remote sensors used by CHWs can provide remote monitoring of ECG (e.g., AliveCor’s KardiaMobile), blood pressure (e.g., AliveCor’s Omron Evolv), glucose and physical activity ^10^. However, large scale data acquisition, pre-processing and validation for intelligent decision-making remain key challenges with existing methods. Sophisticated digital platforms are also being used to facilitate and augment CHW training and supervision ^10,41,42^, to support CHW-CHW communication and collaboration (e.g., virtual informal groups and learning networks ^43,44^ which enable CHWs to exchange information and pose questions to peers ^10,45^), and to provide electronic decision support ^10^. Digitalization is also increasing opportunities for data collection and analysis ^10^ and for outcome evaluation of CHW-delivered interventions, thereby increasing opportunities for key CHW-focused policy advocacy.

### 3.2 Limitations of the Current Approaches

The increasing use of digital technologies among CHWs is demonstrating the potential to expand the CHW footprint; however, *existing digital technologies incorporated into CHW practice remain ad hoc and lack integration*. While alerts, notifications, and reminders facilitate adherence to protocols and improve quality of CHW-provided care ^10^, the current CCC model remains limited by a lack of ubiquitous, closed-loop monitoring and intervention, thereby limiting CHWs to a passive, episodic and reactive approach to client monitoring. CHWs typically provide infrequent (e.g., monthly) home visits and rely on their own episodic observations as well as on client-reported symptoms to inform care and/or referral decisions. However, direct CHW observation and client reporting of symptoms both reveal only an episodic snapshot of a client’s physical and mental health and lifestyle. Moreover, under the current CCC model, the CHW must rely upon the client and/or the client’s caregiver (typically a family member) to be accurate reporters of client health symptoms, both in the present moment of CHW assessment as well over the period of time between check-ins ^46^. CHWs must rapidly synthesize their own observations with the information they receive from the client and/or caregiver in order to decide an appropriate course of action (e.g., re-check, refer, call or immediate medical assistance). This places a large burden of responsibility on the CHW (who must synthesize a large amount of information in a short span of time) as well as on the client and/or caregiver (who must accurately and honestly appraise and report their experiences to the CHW) ^46^. To improve upon this current model of care, and to maximally amplify and expand the CHW footprint, a novel integrated, intelligent and automated closed-loop platform designed for, and with, CHWs is critically needed.

## 4. A New Model: Digital Health-Enabled Community-Centered Care (D-CCC)

We propose D-CCC as an improved model to move CCC into the future. D-CCC is a novel integrated, intelligent and automated closed-loop technology platform designed for, and with, CHWs. By targeting the human-technology partnership at the level of the CHW, D-CCC amplifies human connection and collaboration and maximally expands the CHW footprint through a digitally connected and empowered CHW workforce. By including smart development of personal models unique to each individual client, holistically represented by a high-dimensional cover of multiple knowledge layers, D-CCC additionally amplifies the level of contextual and individualized care CHWs are able to deliver.

### 4.1 The D-CCC Model

The D-CCC model transforms the manual and restricted aspects of CHW work into a scalable, digital and intelligently automated space. By expanding CHW-client communication, as well as CHW collaboration, supervision and support, D-CCC may increase the quality of services delivered, in terms of personalization, cultural appropriateness, and timeliness. Through smart supervision, D-CCC enables CCC organizations to employ and supervise a larger group of CHWs with the same number of CHW supervisors. The D-CCC model also expands the CHW footprint by enabling CHWs to serve a larger volume of clients and/or to provide more services to each client *without increasing undue burden on this vital frontline workforce*. Through automation of manual tasks and critically connecting CHWs to clients, other CHWs, supervisors and ongoing education/training, D-CCC may also empower CHWs and improve worker experience.

Figure 1(a) illustrates the current CCC model, which is limited by unilateral communication, poor scalability and lack of digitization. In comparison, Figure 1(b) illustrates how the D-CCC model addresses these limitations and amplifies the CHW footprint by increasing CHW-client, CHW-CHW and CHW-supervisor communication and collaboration. D-CCC is a human-technology partnership which integrates CHWs, CHW supervisors and communities through a scalable digital medium which empowers CHWs and improves efficiency and quality of care delivery. Through an intelligent and scalable AI loop, D-CCC brings critical stakeholders under a unified communication, collaboration, education/training and care delivery model.

### 4.2 D-CCC’s AI-Enabled Closed-Loop Health Platform

CHWs occupy a variety of roles within communities and CHW-delivered interventions and target outcomes vary according to client populations served. Consequently, a ‘one-size-fits-all’ platform design will not be successful in empowering this uniquely diverse frontline workforce. D-CCC design is therefore modular and adaptable and may flexibly translate to different languages spoken in local communities. Additionally, as cultural and contextual needs will vary according to populations served, D-CCC is designed to be portable across diverse communities. The D-CCC platform caters to different stakeholders viz., clients, CHWs and registered providers (RPs), such as registered nurses (RNs). Further, the D-CCC platform handles diverse aspects such as communication between stakeholders, data collection and analysis, training and education, and autonomous and intelligent decision-making.

The D-CCC platform design is built on the following modules:

1. Interactive Stakeholders (i.e., Smart Communication): This module comprises the stakeholders (i.e., clients, CHWs and CHW supervisors [RPs]) who are interacting through different communication channels viz., client-CHW, CHW-CHW, and CHW-RP.
2. Monitoring: This module focuses on continuous monitoring and updating of physiological and contextual information collected subjectively and objectively.
3. Health Estimation: This module focuses on analyzing the data collected from clients (through the monitor module, CHWs, medical history, etc.) to determine client health status in real time.
4. Knowledge Base: This module manages the storage, access, and retrieval of knowledge built based on the data collected from clients, CHWs, and RPs to enable personalized model building.
5. Personal Models: This module builds cognitive learning models from client physiological and contextual data to provide autonomous intelligent decisions for interventions that are individually tailored for each client.
6. Smart Recommendation: This module autonomously predicts appropriate healthcare and lifestyle recommendations for clients leveraging their personalized models. Recommendations are delivered to CHWs or directly to clients depending on nature, context and risk-level.
7. Smart Supervision: This module supports the RPs in supervising CHWs in specific areas that are determined through automating repetitive tasks and reflecting on personal models and knowledge base.
8. Smart Assistance: This module improves quality of healthcare services by supporting CHWs making intervention procedural decisions in the field.
9. Smart Training: This module provides training for CHWs to learn new digital technologies and enables virtual training modules to augment traditional, in-person CHW training methods.

As shown in Figure 2, our AI-enabled D-CCC platform critically integrates these modules to create a holistic and data-driven approach which connects clients, CHWs and supervisors in a continuous loop of measurement, estimation, guidance and influence.

**Figure 2:**
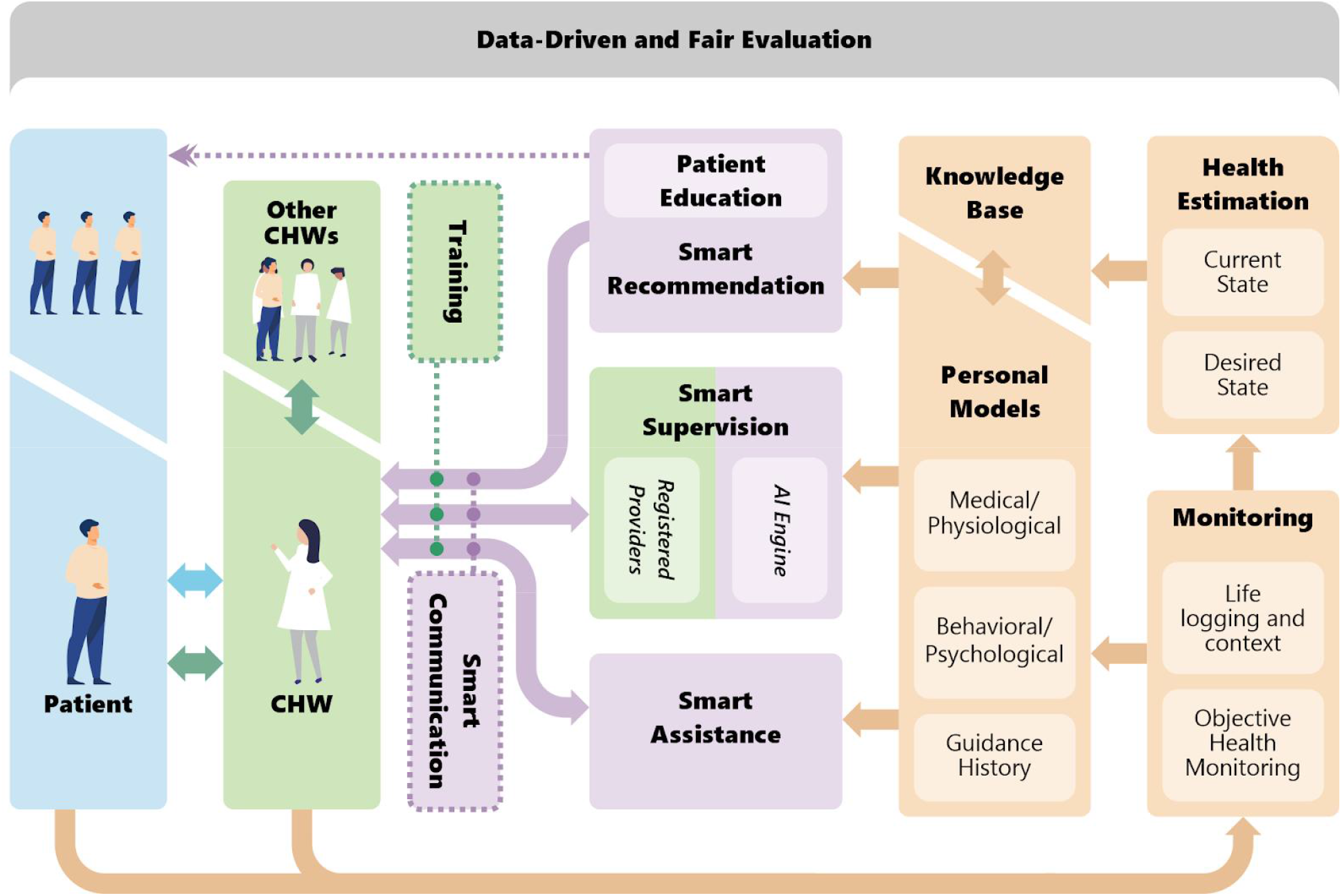
AI-enabled cybernetic platform to empower CHWs of the future.

#### 4.2.1. Interactive Stakeholders

The primary emphasis of the D-CCC model is to enable an interactive multi-way communication among different stakeholders involved in health-related service delivery networks. Clients within the community, CHWs, and CHW supervisors together form the key stakeholders. We focus on establishing digital communication channels in a scalable manner to establish and maintain active communication among all stakeholders. Figure 2 shows the different combinations of multi-way communication channels among clients, CHWs, and supervisors. We classify communication into the following channels.

##### Client-CHW

This is the point of communication between clients and CHWs. As a baseline, each client is assigned a CHW who is responsible for providing health-related services. Traditionally, the client-CHW interaction is scheduled over fixed time windows and/or on-demand subject to the client’s needs. D-CCC transforms this to a continuous interaction and the digital communication medium is available to all clients served.

##### CHW-supervisor

This is the point of communication between CHWs and their supervisors. Each CHW has a supervisor assigned for supervision and feedback. As service delivery demands change, CHW supervision can be extended to include additional supervisors (RPs) with expertise regarding different areas of healthcare. CHW supervision is transformed to an on-demand, real-time interaction through the digital medium.

##### CHW-CHW

This is the point of communication between different CHWs. Communication and collaboration among CHWs promotes knowledge transfer and exchange of insightful information regarding interventions which may be adapted to different community-specific challenges as needed.

#### 4.2.2. Monitoring

The Monitoring module is responsible for continuous monitoring of client physiological signs, contextual information, life-log, surveys, and ecological momentary assessments (EMAs). State-of-the-art wearable (e.g., smart rings, watches, patches), portable (e.g., smart EKGs, stethoscopes, blood pressure monitors), and stationary sensors (e.g., smart beds, fall detection cameras) as well as mHealth (smartphone-based) solutions enable such ubiquitous monitoring. Context and lifelogs are recognized using contemporary life event recognition technologies such as situation specific recognition (e.g., smart home), computer vision-based recognition (e.g., surveillance or wearable cameras), and sensor-based recognition (e.g., accelerometer, and GPS). These enable capturing of low-level lifelogs (e.g., step count, GPS, venue, or physical activity) as well as high-level daily activities (e.g., commuting, shopping, socializing) that enable assessment of an individual’s lifestyle. This hyper dimensional sensory data is dedicated to determining a quantifiable state of health of a given client. Coupling physiological signs with contextual data validates the perspective of objective health monitoring complementing the subjective data collected through self-reports and EMAs.

#### 4.2.3. Health Estimation

The Health Estimation module is responsible for determining the health status of the client. The high-dimensional and holistic information collected by the Monitoring module can be used to identify the client health state. Each client is unique and presents multiple possibilities of health states. A key functionality of the Health Estimation module is to process different modalities in order to properly estimate health variables in different dimensions. For each client, this module holds a health status that is deemed to be safe, termed as the client’s ‘desired state.’ The Health Estimation module estimates the current state (i.e., client’s current health status) and compares the current state to the desired state to estimate the overall health safety of the client. The health status as determined by the Health Estimation module assists in building personal models of each client and guides intervention decision-making made by the CHWs, supervisors, and recommendation engines.

#### 4.2.4. Knowledge Base

The Knowledge Base module consists of CCC-related facts, information, and skills acquired through past experience of different stakeholders (e.g., CHWs, supervisors [RPs], clients, etc.) as well as theoretical or practical knowledge available in the field of CCC. This module is a dynamic component in the sense that it can continuously learn and update its knowledge based on new inputs extracted from the data generated by the entire pool of clients, CHWs, and supervisors. Each client’s data includes physiological parameters monitored continuously, contextual information that is associated with each instance of lifelogging, as well as intervention procedures performed in the past and health estimation logs. Each CHW’s data includes the list of assigned clients, log of intervention performed per each client, effectiveness of the interventions, as well as supervision and training provided by supervisors. The Knowledge Base module guides building of learning analytics and cognitive models in the subsequent modules that make personalized intelligent decisions.

#### 4.2.5. Personal Models

The Personal Models module focuses on building analytical models for intelligent decision-making tailored to a specific individual. That every individual constitutes a biological system which responds differently to different inputs forms the basis for the P4 (predictive, preventive, personalized, and participatory) ^47^ approach to healthcare. To build an approach that makes P4 a persistent action to guide CHWs and clients, it is important to estimate and build a personal model of each client. In many areas relevant to personalization, a model is built by collecting data in the context of the application. Google, Facebook, Amazon, and Netflix are used as prime examples of personalized recommendations using models built for each individual when they interact with different applications on their platforms. The Personal Models module relies on the physiological parameters, context, and health status data of each client, collected by the Monitoring and Health Estimation modules. A variety of methods such as machine learning can be used for data analysis and cognitive modeling for autonomous decision-making. As shown in Figure 2, we divide the personalized modeling into the sub-modules of medical profiling, behavioral profiling, and guidance history.

The Medical and Physiological Profile sub-module builds a personalized medical model for each client from the physiological data and health status estimate. This creates a baseline for each client’s health status and provides an intuition on determining the relative health safety under the current conditions. For example, a specific health status of client A can be perceived more alarming as compared to client B, who has a history of the same condition.

The Behavioral and Psychological Profile sub-module builds a personalized behavioral model for each client from contextual data. This creates an insightful perspective on each client’s health status and their response to interventions under specific circumstances. For example, two clients A and B could have the same health status but different contexts, which calls for different intervention procedures. This allows CHWs to make decisions that are customized to both the client’s physiological status and their context.

The Guidance History sub-module logs the past health conditions of a client and their contexts, the responsive intervention procedures imposed under each context, and the response of the client to each intervention use case. This allows for development of a baseline understanding for CHWs to decide on appropriate intervention in the current scenario. Given the history of interaction and guidance provided by CHWs in the past under similar situations, decisions on an improvised intervention or re-using a previously successful intervention can be made.

#### 4.2.6. Smart Recommendation

The Smart Recommendation module focuses on delivering fully autonomous health recommendations to clients and to CHWs based on the risk-level and type of a recommendation. We combine each client’s physiological and contextual data from the Knowledge Base and Personal Models modules to build smart recommendation systems. Smart recommendation is a proactive strategy to promote client self-management and improve quality of life, particularly in less acute scenarios (e.g., recommendations related to diet, physical activity, sleep, stress, medication reminders). Smart recommendations sent out to clients target everyday health maintenance and education regarding healthy life choices. The Smart Recommendation module infers to the cognitive and analytical models from the Personal Models module to make autonomous recommendations. The autonomous recommender primarily aims to bridge the gap between the increasing number of clients in need and the relatively lower number of CHWs and RPs and is based upon acuity of client condition. Recommendations are also sent out to the responsible CHW to synchronize the information flow. The decision of whether the recommendation needs to be delivered directly to the client or administered through the CHW depends on the settings and recommendation type. For instance, an elderly population with less access or tendency to use smartphones may prefer to receive recommendations through their CHWs while younger technology users such as pregnant mothers might be willing to directly and continuously receive health promotion recommendations. The Smart Recommendation module thus adds a layer of cyber-healthcare service delivery to guarantee safe and timely intervention for all clients.

#### 4.2.7. Smart Supervision

The Smart Supervision module automates the workflow between supervisors (RPs) and CHWs to improve supervision efficiency both qualitatively and quantitatively. While CHWs may receive appropriate training beforehand, there is a need for continuous interaction between CHWs and supervisors for evaluation, feedback and support in a supervisory capacity. In the CCC model, supervisors manually organize supervisory tasks with different CHWs, consuming more time, at a higher cost, and lower productivity. D-CCC’s Smart Supervision module directly addresses these limitations with automated supervision support using AI models. Each CHW handling multiple clients in different contexts presents each RP with different supervisory challenges. The Personal Models and Knowledge Base modules contain cognitive analytical models about different clients. The Smart Supervision module integrates all these personalized models into an AI engine that serves as the oracle to the supervisors. This allows supervisors to narrow down the focus to specific issues that an individual CHW may have, given the collective record of clients specific to that individual CHW. With a common and more complete understanding of the client’s context, the interaction between RPs and CHWs becomes significantly qualitative and reduces the time overhead. Supervisors can also automate trivial repetitive tasks allowing them to supervise a larger number of CHWs, thereby improving overall productivity of care delivery. Multi-way communication among different CHWs and supervisors additionally allows for a range of choices to match the supervision requirements of the CHWs with expertise of the RPs. Such holistic interaction enables scalability of the supervision strategy.

#### 4.2.8. Smart Assistance

The Smart Assistance module supports CHWs to deliver continuous high quality healthcare services in the field, particularly in scenarios with limited access to resources. Applying intervention procedural knowledge for clients with diverse contexts can be challenging for CHWs in the field. The Smart Assistance module supports CHWs to make client-specific decisions using personalized models. Smart assistance is a relatively reactive and on-demand service that is intended to handle acute scenarios. It enhances the eventual decision-making of the CHWs in the field when they do not have immediate access to supervision. One approach is to implement customized chatbots that interact with CHWs in real time to provide appropriate decisions regarding client care. This becomes extremely useful when supervisors may not be accessible and/or when CHWs encounter a challenging situation with which they have limited expertise. The chatbots employ natural language processing to parse the text information provided by CHWs to make sense of the client care situation. The Smart Assistance module accesses the client’s medical and physiological profile, behavioral and psychological profile, and previous history of healthcare assistance provided from the knowledge base. Upon identifying the specific client, the Smart Assistance module infers the personalized model of the client to make autonomous decisions on necessary health-related services to be provided. The chatbot interactively serves as a virtual supervisor to support CHWs with instructive intervention procedures.

#### 4.2.9. Smart Training

The Smart Training module provides training for CHWs to learn new digital technologies and augments traditional methods of CHW training for smart and connected virtual and/or hybrid learning methods. Key challenges identified in the literature regarding current use of digital technologies include CHW digital health literacy ^10^ and the need for effective training for CHWs to adopt digital tools ^5^. The Smart Training module integrates ongoing CHW digital health training and provides CHWs with an automated technology ‘help desk’ for additional assistance as needed. The Smart Training module additionally augments traditional, in-person CHW training by providing ongoing virtual case-based learning modules, which may be assigned by CHW supervisors and completed by CHWs in an independent and self-paced manner.

## 5. Case Study: Smart, Connected, and Coordinated Maternal Care for Underserved Communities (UNITE)

The D-CCC model is currently being piloted with our community partner site, MOMS Orange County ^48^ (MOMS OC), and we present this as a case study below.

RN home visits for at-risk pregnant women in Orange County (OC) California became infeasible in the early 1990s due to the county’s financial constraints. In 1992, in response to the county’s maternity care crisis, a nonprofit organization MOMS OC ^48^ was founded to implement a CCC model (Figure 3) for maternal care (MC) delivery. MOMS’ CCC model is a CHW-delivered care-coordination and home visitation (HV) program wherein RNs supervise and train CHWs, who in turn provide culturally and linguistically appropriate services, conduct HVs and deliver group education to the low-to-moderate risk women and families served by MOMS.

**Figure 3.**
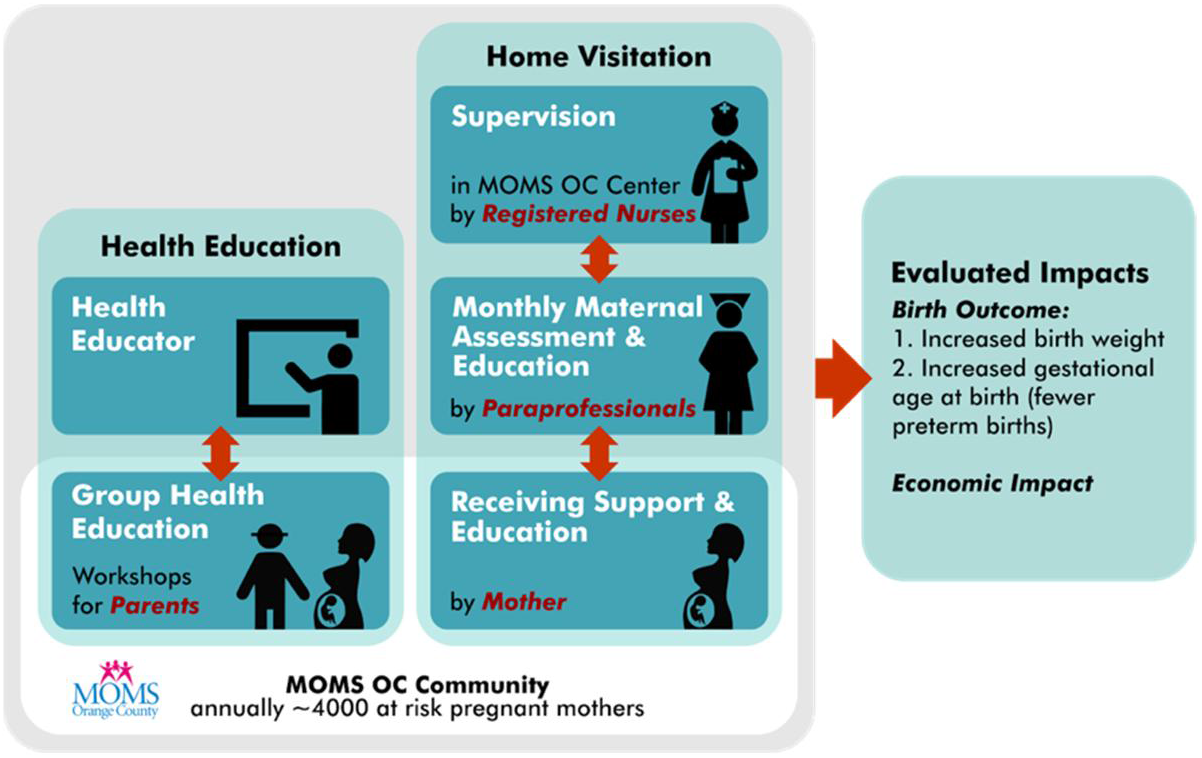
MOMS’ CCC Model.

The MOMS CCC model has demonstrated cost-efficiency ^49^ and improved birth outcomes ^50^. However, it remains limited by barriers and gaps common to CCC models generally and which present opportunities for incorporation of technology. These include: (1) *Limitations regarding outreach* using traditional mechanisms (e.g., word of mouth, phone calls, and ads), particularly among disadvantaged communities. This presents opportunities for technology-enabled community outreach; (2) *Limitations regarding timely intervention*, including early MC and education. This presents opportunities for new community-driven intervention enablement assisted by digital and social media; (3) *Limitations regarding monitoring*, as infrequent CHW HVs (typically monthly) and mothers’ self-reported screenings both reveal only a snapshot of each mother’s physical and mental health and lifestyle. This presents opportunities for incorporation of low-cost, non-intrusive wearable IoT infrastructure to enable ubiquitous monitoring; and (4) *Limitations regarding communication between MC providers*, with CHWs typically unable to access health data stored as electronic health records and MC providers typically unable to easily access information collected by CHWs. This presents opportunities for networking, IoT, smart data mining, and community-enhanced recommendation systems.

Identifying opportunities for D-CCC to overcome these gaps and improve the quality of care delivered, we partnered with MOMS to launch UNITE ^51^, a pilot community engagement model for MC that is smart (deploying ubiquitous monitoring and lifelogging), connected (bringing a together a diverse cast of community members including mothers, families, care providers, and outreach resources) and coordinated (using technology to proactively reach out to the community and use personalized intervention and education for the women).

UNITE deploys technology-enhanced community care coordination and education coupled with a human-in-the-loop monitoring-intervention system to: 1) proactively reach out to the (expanded) underserved community, including mothers, families and friends; 2) provide valuable personalized and community-enhanced information for CHWs and RNs to better support clients and tailor per individual needs; and 3) combine fine-grained and personalized information together with monitoring/intervention and community outreach/education to promote healthier lifestyle for pregnant mothers through self-management. UNITE leverages emerging technologies such as Wearable Internet-of-Things (WIoT), community-enhanced learning and recommendation, big data analytics, context recognition, lifelogging and social media, as well as a multi-disciplinary partnership to bring connectivity and smartness to MOMS’ existing CCC model. UNITE also brings smartness to MOMS’ CCC model via ubiquitous monitoring, open information sharing, and community-enhanced personalized intervention and education. UNITE additionally strengthens connectivity among stakeholders and improves connection and coordination across providers and agencies focused on prenatal and postnatal health.

Through lifelogging, context recognition, and health monitoring, UNITE builds a holistic digital phenotype of enrolled mothers using multi-modal data capture. Smart mining algorithms are designed for cause assessment through personalized models. Maternal self-management is improved and incentivized through personalized community-enhanced recommendation systems, technology-enhanced community care coordination and education. The core components of the UNITE model are ubiquitous monitoring and a recommendation system capable of dynamically supporting a healthy lifestyle of enrolled mothers during and after pregnancy. Our guiding design principle for UNITE was recognizing how community-specific factors pertaining to each mother enhance individual monitoring and interventions and enable more personalized recommendations to motivate better self-management. UNITE thus integrates culture- and context-sensitive mechanisms to enhance technology acceptance for improved maternal self-management and enhances connection and coordination with CHWs and other care providers and agencies focused on improving pre- and postnatal maternal health.

As part of the UNITE project, a proof-of-concept version of the AI-enabled closed loop system has been implemented to enable smart community-centered maternal care. We designed a Wearable Internet-of-Things (WIoT)-based health monitoring flexible and adaptable platform to enable seamless capture of sensory data streams (using wearables and smartphones) and self-reported data from enrolled mothers (using questionnaires and EMA). UNITE’s dashboard allows for interaction between CHWs and enrolled mothers, provides visualization and interaction with the backend system, and performs sophisticated analytics and data mining to build a holistic personal model of each enrolled mother. Each enrolled mother’s personal model couples physical and psychosocial data capture to generate feedback and recommendations to enrolled mothers, CHWs, CHW supervisors and other health providers with the goal of promoting healthy pregnancy outcomes.

An example intervention delivered through the UNITE platform is a smart intervention designed for MC called the Two Happy Hearts (THH) program ^52^. Objectives of THH include enhancing mothers’ awareness of their emotional states and empowering them to reduce their stress through an integrative health approach which includes mindful breathing, home-based exercise and health coaching delivered by CHWs. Key components of THH include personalization of the mindful breathing and strength movement components specific to the individual health state of each mother as well as enabling CHW-delivered health coaching through virtual visits to enhance mothers’ self-management skills. THH combines continuous measurements (e.g., heart rate (HR), heart rate variability (HRV), respiration rate (RR), skin temperature, sleep, and physical activity) collected using wearable sensors and lifelogging and high-level daily activities (e.g., working, commuting, shopping, or relaxing) collected using a passive smartphone app to build personal models and provide individualized smart recommendations. For example, a mother may be recommended to use our mHealth-based mindful breathing app when a stressful moment is captured through real-time analysis of her physiology and environmental context. Another example of an intervention delivered through the UNITE platform’s closed-loop system is a home-based safe exercise program which is overseen remotely by CHWs. This evidence-based exercise program, designed using the pregnancy and postpartum exercise guidelines provided by the American College of Obstetricians and Gynecologists (ACOG), includes strength movement, aerobic movement and mindful breathing. UNITE’s real-time monitoring of intensity and duration of exercise using wearables (e.g., by measuring HR elevation, VO2 Max, RR, and ratings of perceived exertion) critically enables individualization of the intervention as well as safety monitoring throughout the exercise program.

## 6. Discussion

The current CCC model limits CHWs to a passive, episodic and reactive approach to client monitoring which is reliant upon accurate reporting from clients and/or client caregivers. In contrast, D-CCC represents a pioneering vision for the future of CCC which has the potential to transform the current state of manual, episodic and restricted CHW-delivered services into an expanded, digitally interconnected, collaborative and empowered community health workforce of the future.

D-CCC’s platform design expands upon our pilot project UNITE. While the UNITE model targets CHW-delivered MC to underserved mothers in Orange County, California, D-CCC is designed to be scalable and portable across diverse communities. Furthermore, while UNITE was designed to leverage technology to facilitate client self-management, D-CCC instead targets the human-technology partnership at the level of the CHW. By amplifying human connection and collaboration through an empowered and digitally connected CHW workforce, D-CCC is designed to amplify CHWs’ reach and enhance diffusion of CHW-delivered health information within communities ^5,36,37^. D-CCC is also designed to support CHW-delivered health-related interventions among client populations less able to engage in self-management and/or demonstrate facility with technology (e.g., older adults with dementia). As CHWs occupy a variety of roles within communities and as CHW-delivered interventions and target outcomes may vary widely according to client populations served, a ‘one-size-fits-all’ platform design will not be successful in empowering this uniquely diverse frontline workforce. D-CCC’s modular design critically allows for flexibility, scalability and portability across CHW-delivered interventions in accord with health outcome targets, language of delivery and diversity of client populations served.

D-CCC critically brings all stakeholders under a unified communication, collaboration, education/training and care delivery model. D-CCC’s modular, AI-enabled algorithm-generated data loops will provide CHWs with continuous, real-time and actionable biometric data individualized to each client. This data will provide CHWs with client data trends, alerts and guidance in real time and can be used to notify CHWs of client health status and provide feedback and guidance to clients to help them attain their individualized optimal health state. Each client’s unique data structure will contain health monitoring schedules, client vital signs, a history of allocated CHWs, recommended healthcare actions and feedback from CHW supervisors. This data has the potential to improve health-related services by providing CHWs with an efficient means of retrieving client past medical history and determining subsequent course of action. As client data are logged continuously and populate automatically, CHWs will no longer need to input data manually into records or spreadsheets. This may save worker time result in more accurate reporting of client data. CHWs will be alerted when client data fall outside of predetermined parameters and aggregate data trends will be available for review continuously and in real time, allowing CHWs to quickly visualize client health-related data trends over time.

D-CCC’s design addresses key challenges identified in the literature regarding the current use of digital technologies by CHWs ^5,10^, including a lack of evaluation of outcomes, development of digital tools and apps without cultural relevance, lack of access to and/or knowledge of mobile technologies within communities, need for effective training for CHWs to adopt digital tools, inadequate CHW supervision and the need for improved communication between and among health care teams working with CHWs. D-CCC allows for fair and data driven evaluation of CHW-delivered interventions, thereby increasing opportunities for key CHW-focused policy advocacy. Data-driven evaluation of CHW-delivered interventions is key to moving community organizations away from fee-for-service or grant-based governmental support models and toward value-driven models for CHW reimbursement. Data-driven evaluation enables CHW-delivered interventions to be linked to improvements in community health outcomes and/or cost-savings, and will be critical to transforming CCC and making the needed a shift away from a patient-centered medical care system to an ‘upstream’ health promotion approach which meets the health and social needs of individuals in the communities where they live ^1^. However, D-CCC’s data-driven algorithms, which are key to personalized model-building, also mean an unprecedented scope of sensitive data collection, and the digital health ecosystem presents new ethical challenges and considerations, particularly in vulnerable populations. A D-CCC-based platform needs to critically protect data privacy and security through methods such as end-to-end encryption and distributed security schemes (e.g., blockchain technology) and proper data governance policies and tools.

While human-technology partnerships and design of new technologies to augment human performance have incredible potential to empower future workers and to mitigate health inequities, it is important to consider fully the impact of AI on future workers and communities. A vital consideration is identifying potential for inadvertent creation or exacerbation of health inequities, particularly among vulnerable populations. Consideration of socio-cultural aspects *during D-CCC platform design*, and iteratively examining how these affect platform acceptability, feasibility and reliability will be critical to avoid further propagating health inequities ^53^. To this end, D-CCC design needs to occur iteratively and in close collaboration with CHWs, CHW supervisors and clients. This process needs to continue throughout implementation and piloting of D-CCC within the community: as CHWs use D-CCC in the field, regular and ongoing meetings will allow for rapid changes to be made, according to ongoing CHW, supervisor and client feedback.

## 7. Conclusion

The increasing use of digital technologies among CCC organizations is demonstrating the potential to expand the CHW footprint by enhancing CHWs’ reach and diffusion of health information within communities ^5,36,37^. However, existing digital technologies incorporated into CHW practice remain ad hoc and lack integration. We propose D-CCC as a pioneering vision to move CCC forward into the future. D-CCC’s novel integrated, intelligent and automated closed-loop technology platform integrates CHWs, CHW supervisors and communities through a scalable digital medium which empowers CHWs and improves efficiency and quality of care delivery. By targeting the human-technology partnership at the level of the CHW, D-CCC amplifies human connection and collaboration and maximally expands the CHW footprint through a digitally connected and empowered CHW workforce of the future.

## Data Availability

This paper does not include any data.

## Acknowledgments

This research was supported by the U.S. National Science Foundation under the grants FW-HTF 2026614 (“D-CCC: Digital Health for Future of Community-Centered Care”) and SCC 1831918 (UNITE: Smart, Connected, and Coordinated Maternal Care for Underserved Communities). We would also like to acknowledge our collaborators in the UNITE project, in particular, the co-PIs Dr. Yuqing Guo and Dr. Marco Levorato. The authors would like to also thank Arman Anzanpour for his assistance with the conceptualization and visualization of the figures.

